# Transmural side holes in covered metal stents do not reduce bile duct occlusion or cholecystitis: a single center study

**DOI:** 10.64898/2026.05.14.26353216

**Authors:** Alexandra M. Stendahl, Jessica X. Yu, Saad Jazrawi, Emily Jonica, Jennifer Rodriguez, Siddharth Javia, Kaveh Sharzehi, Gregory A. Coté

## Abstract

**Background and Study Aims:** Fully covered, self-expandable metal stents (FCSEMS) are used to treat biliary strictures. FCSEMS with transmural side holes may facilitate cystic duct drainage to mitigate risk of cholecystitis and impact other stent-related adverse events such as migration and occlusion. This study compared rates of premature stent occlusion and acute cholecystitis among patients with biliary strictures who underwent first-time placement of a FCSEMS with or without transmural side holes.

**Patient and Methods:** This was a retrospective cohort study of adults who underwent endoscopic retrograde cholangiopancreatography (ERCP) with FCSEMS between April 2022 to April 2025 for malignant or benign extrahepatic bile duct strictures. The primary outcome was premature bile duct occlusion. The secondary outcome was acute cholecystitis among patients with an intact gallbladder.

**Results:** Among 219 patients meeting enrollment criteria, 57 (26%) had side holes. The rate of premature stent occlusion was similar with transmural side holes (12%) vs. without (11%, HR 1.02, 95% CI 0.42–2.43, p = 0.96). Among patients with an intact gallbladder (n=129), acute cholecystitis rates were similar with side holes (6%) or without (4.8%, HR 1.01, 95% CI 0.22–4.5, p = 0.99).

**Conclusions:** In this preliminary single center retrospective study, FCSEMS stents with side holes did not reduce rates of premature bile duct stent occlusion or acute cholecystitis compared to FCSEMS without side holes.

## Introduction

Endoscopic retrograde cholangiopancreatography (ERCP) with placement of a bile duct stent remains the first-line approach to the palliation of extrahepatic bile duct obstruction secondary to cancer or benign etiologies. Given their larger diameter and need for fewer reinterventions, self-expandable metallic stents are generally preferred over plastic stents for malignant and most benign etiologies. Fully covered self-expandable metal stents (FCSEMS) are increasingly preferred over uncovered varieties because of their removability, an especially important characteristic for benign or indeterminate etiologies.^1–5^ While FCSEMS confer lower rates of tissue ingrowth, patency rates are often similar to uncovered SEMS because of higher rates of spontaneous stent migration or clogging with debris.^3,6–8^ FCSEMS with anti-migration anchoring fins were developed to address the issue of spontaneous stent migration.^9^

Acute cholecystitis is a known risk of ERCP, but previous studies suggest metallic stents confer a higher rate due to their radial force and secondary compression of the cystic duct orifice; the rate may be highest with FCSEMS which should prevent drainage through the interstices of the stent (**figure 1**).^10^ In addition to stent placement, other risk factors for post-ERCP cholecystitis include opacification of the gallbladder, stenting across the cystic duct orifice, distal malignant biliary obstruction, tumor involvement in the cystic duct, and gallbladder stones; rates of cholecystitis are 4-15% following FCSEMS placement.^2,4,10–14^

**Figure 1:**
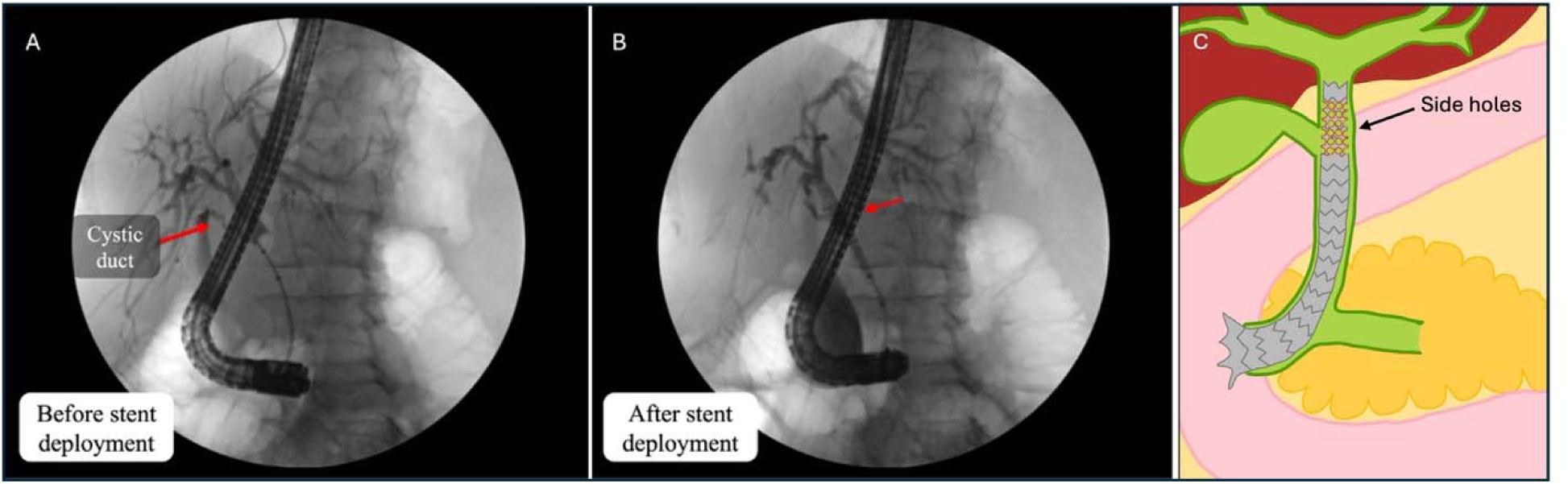
Visualization of the cystic duct during ERCP. (A) Cholangiogram demonstrates insertion of the cystic duct located immediately above the proximal margin of the bile duct stricture. (B) A fully covered metal stent with side holes is deployed, with the proximal aspect of the stent crossing the cystic duct insertion, with the red arrow highlighting a radiopaque band in the region of the transmural side holes. (C) A schematic of a stent with proximal side holes allowing for drainage of the cystic duct.

A hybrid SEMS with holes throughout the plastic sheath (designed to reduce migration by allowing epithelial ingrowth) has also shown mild reduction in recurrent biliary obstruction, attributed in part to reduced migration, but may additionally preserve drainage through the stent. [6], [7]

In parallel, newer stent designs have included drainage side holes clustered at the proximal end of the FCSEMS. In theory, this allows for cystic duct drainage while preserving the benefits of a FCSEMSm however cholecystitis and biliary occlusion outcomes remain uncharacterized.

This study compares outcomes between FCSEMS with and without drainage side holes in a retrospective, single center pilot study to inform future larger multi-center studies.

## Methods

This was a retrospective cohort study of adult patients undergoing ERCP with placement of a FCSEMS with proximal and distal anchoring fins (GORE^®^ VIABIL^®^ Biliary Endoprosthesis, W.L. Gore & Associates, Flagstaff, Arizona, USA) between April, 2022 and April, 2025 at a single pancreatobiliary endoscopy referral center (Oregon Health & Science University, Portland, Oregon, USA). The institutional review board at Oregon Health & Science University approved this investigation before commencement of data collection.

All patients ≥ 18 years of age who underwent ERCP with placement of a FCSEMS for the indication of benign or malignant stricture were considered for enrollment. Based on the preference of the investigators, all FCSEMS during the study period were derived from a single manufacturer (VIABIL^®^) and included anti-migration anchoring fins. Patients were excluded if the stent was placed for strictures involving the hepatic confluence (i.e., Bismuth type II-IV) or for non-stricture indications such as post-sphincterotomy bleeding or a postoperative bile leak. Additional exclusion criteria included previous placement of a FCSEMS within the study period or patients lacking follow-up in the medical record.

Eligible subjects were followed through medical record review until planned stent removal (if applicable), surgical resection, death, or until a stent-related adverse event occurred. Otherwise, patients were followed through the medical record until data collection was terminated (January 20, 2026).

The primary outcome was development of premature bile duct obstruction with or without acute cholangitis. Bile duct obstruction was defined by the development of one or more of the following: jaundice, rising liver chemistries without an alternate etiology, abdominal pain with radiographic evidence of stent occlusion (e.g., absence of pneumobilia or presence of tissue ingrowth), or acute cholangitis. The development of acute cholangitis was defined using the 2018 Tokyo guideline.^17^ Acute cholecystitis was not included in this outcome.

The secondary outcome was development of acute cholecystitis, which was restricted to patients with an intact gallbladder at the time of index ERCP. Acute cholecystitis was defined as a perforated gallbladder on imaging, or the combination of consistent symptoms in conjunction with a positive Murphy’s sign and radiographic changes consistent with cholecystitis (i.e., gallbladder wall thickening, pericholecystic fluid, or pericholecystic fat stranding). Fluoroscopy images taken during ERCP were analyzed to assess whether the cystic duct was visible, whether strictures involved the cystic duct, and whether stents visibly crossed the cystic duct. Two of the authors (KS and GAC) assessed the images and independently scored them. In instances of disagreement, a third author (JY) independently assessed the images.

### Statistical analysis

We compared outcomes between patients who underwent ERCP and FCSEMS placement with and without transmural side holes. All analyses were performed using R Statistical Software (v 4.5.0; R Core Team 2026). Cox proportional hazards tests were used to compare time-dependent outcomes. When analyzing the secondary outcome of cholecystitis, we limited the analysis to patients with an intact gallbladder at the time of stent placement. Cox proportional hazards multivariate regression models were used to evaluate the association between patient (age, malignant stricture), stent (presence of side holes, diameter, length >8cm), and duct characteristics (presence of gallbladder, opacification of cystic duct during index ERCP) with the development of premature bile duct stent occlusion (primary outcome) and acute cholecystitis (secondary outcome), using p ≤ 0.10 by univariate testing as the criterion for inclusion.

## Results

During the three-year study period, 303 patients underwent ERCP with placement of a FCSEMS. Of these, 84 (27.7%) were excluded due to a non-stricture indication or lack of follow-up, leaving 219 (72%) for inclusion. Of these, 57 (26%) had side holes and 162 (74%) did not (**table 1**). Patients were followed for a median of 122 days after FCSEMS placement (interquartile range: 45-188). Patients with an intact gallbladder (n=129, 59%) were more likely to receive a FCSEMS with side holes (82% vs. 51%, p<0.001) and have the cystic duct opacified (70% vs. 32%, p < 0.001). Patient demographics and stricture etiology were similar between groups.

**Table 1.**
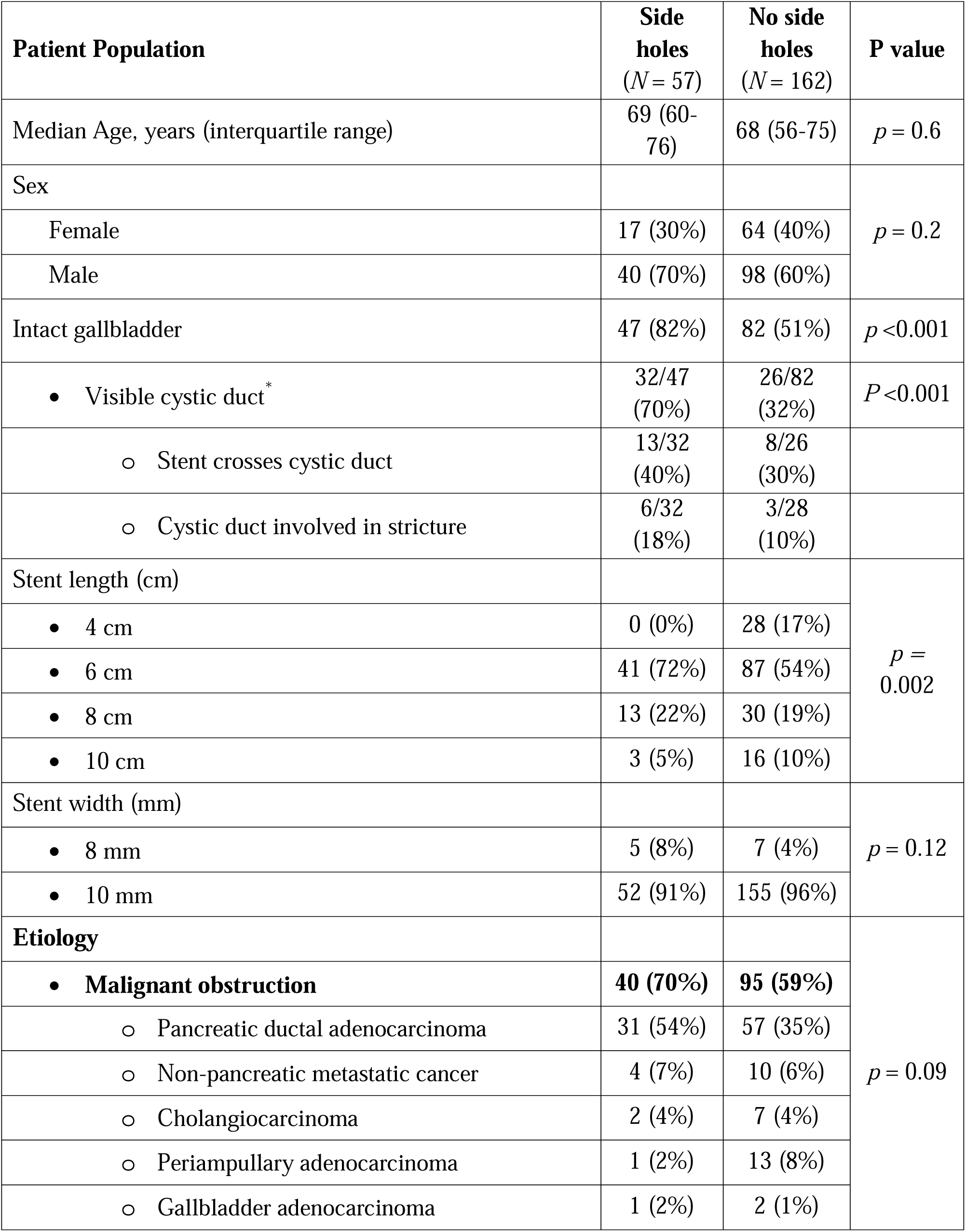
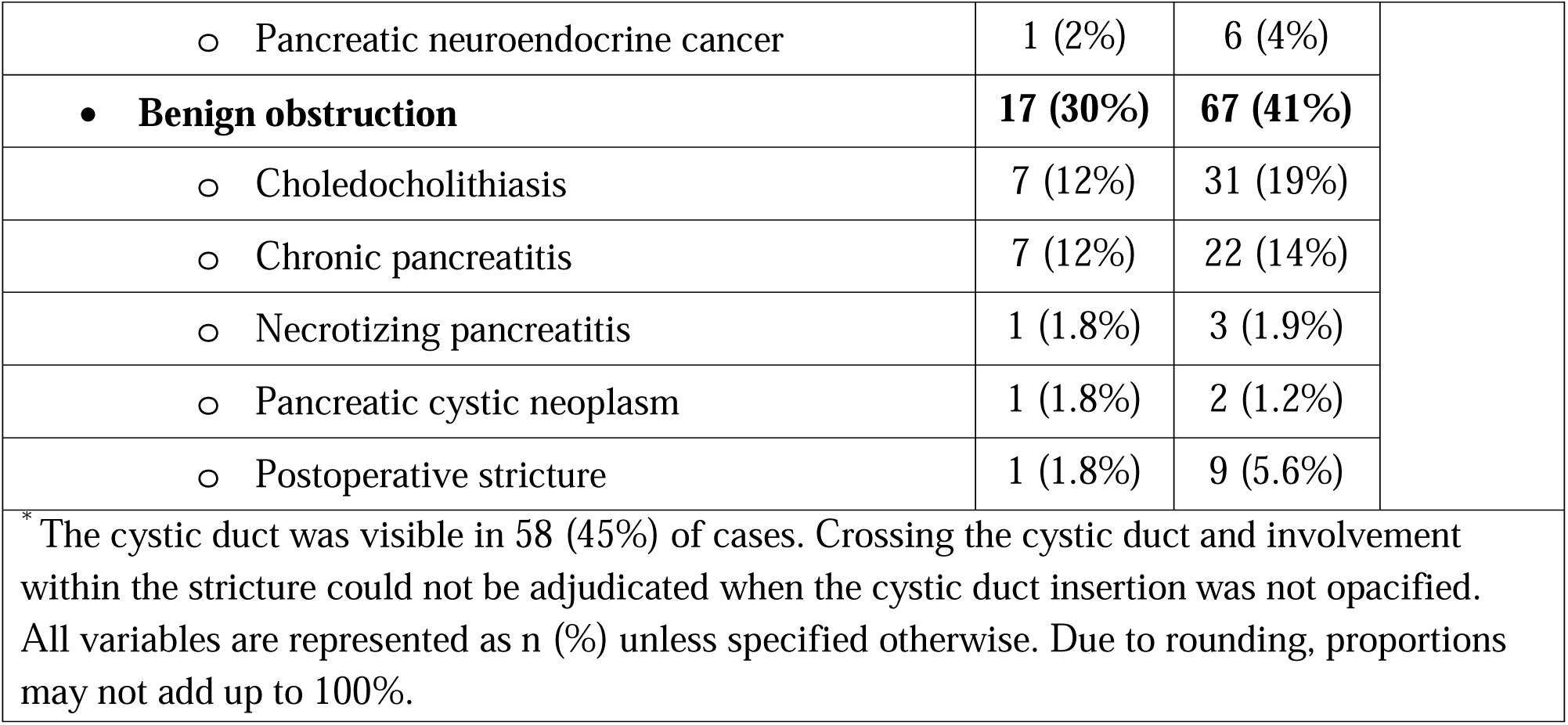
Baseline characteristics.

### Bile duct obstruction

After receiving a FCSEMS with side holes, 7 patients (7/57, 12%) experienced biliary obstruction including acute cholangitis (4/57, 7%), with median time to event 128 days (IQR: 83-166 days). Comparatively, 19 patients (19/162, 11%) who received FCSEMS without side holes experienced biliary obstruction including acute cholangitis (univariate HR = 1.02, 95% CI 0.43-2.44, p=0.96). **Figure 2a** demonstrates Kaplan Meier curve for patients experiencing biliary obstruction.

**Figure 2.**
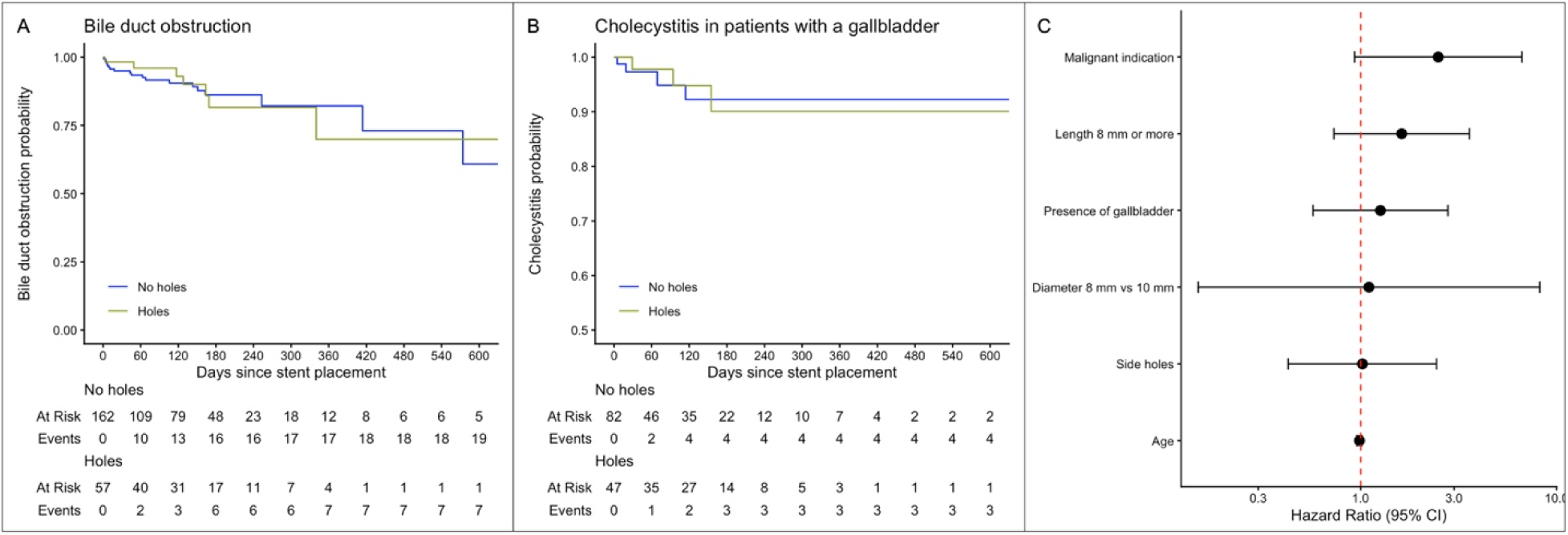
Development of bile duct obstruction or acute cholecystitis Shown are Kaplan-Meier estimates of the proportion of patients who developed premature bile duct obstruction (A) and acute cholecystitis (B). The color bands represent the 95% confidence interval. The median follow-up duration was 130 days (IQR 45-188) for patients receiving a FCSEMS with side holes and 109 days (IQR 43-186) months following FCSEMS without side holes. (C) demonstrates univariate hazard ratios of patient characteristics against the development of premature biliary obstruction.

Table 2 summarizes the results of cox proportional regression analysis of factors potentially associated with an increased risk of biliary obstruction. The presence of holes in the stent was not a significant contributor (HR 1.02, p = 0.96). Malignant bile duct stricture (compared to benign strictures) had a high hazard ratio (HR 2.47, p = 0.07). In multivariate analysis, side holes were not significant, when controlling for malignant vs benign stricture (HR 0.93, p = 0.88).

**Table 2:**
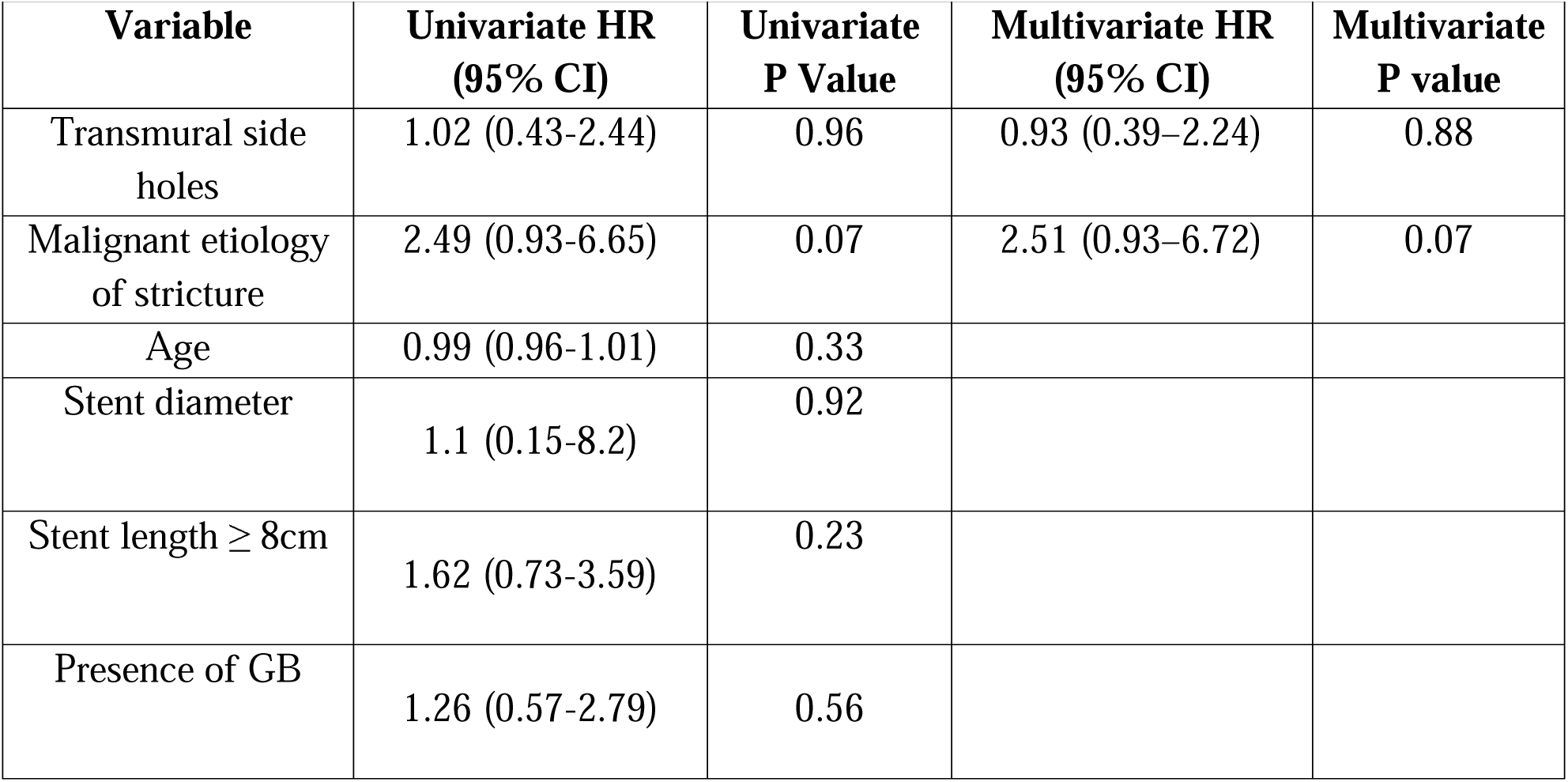
Factors associated with development of premature bile duct obstruction.

### Acute Cholecystitis

Among patients with a gallbladder, all instances of acute cholecystitis (n=7, 5.4%) were among patients with a malignant stricture. After receiving a FCSEMS with side holes, 3 patients (3/47, 6%) experienced cholecystitis, with time to cholecystitis 29, 94, and 155 days for each respective patient. Comparatively, 4 patients (4/84, 4.7%) experienced cholecystitis after receiving FCSEMS without side holes, with time to cholecystitis 5, 19, 69, 114 days for each patient; (HR 0.99, 95% CI 0.22–4.46, p = 0.99). **Figure 2b** demonstrates Kaplan Meier curve for patients with gallbladders experiencing cholecystitis. In evaluating other variables potentially associated with cholecystitis (patient age, stent diameter, stent length ≥8cm, cystic duct opacification on index ERCP), none were found to be associated (p<0.10), so a multivariate model was not constructed (data not shown).

In ERCPs where the cystic duct was opacified (N=58) 21 stents (36%) crossed the cystic duct. 9 patients (15%) had strictures which involved the cystic duct. In patients whose stents crossed the cystic duct, the rate of biliary obstruction was 14% (3/21), compared to 9% (3/32) in stents that did not cross the cystic duct. In patients whose stricture involved the cystic duct, the rate of biliary obstruction was 11% (1/9), compared to 11% (5/47) in patients whose stricture did not involve the cystic duct. Similarly, the rate of cholecystitis in patients whose stents crossed the cystic duct was 14% (3/21), compared to 3% (1/32) in stents that did not cross the cystic duct. In patients whose stricture involved the cystic duct, the rate of cholecystitis was 22% (2/9), compared to 4% (2/47) in patients whose stricture did not involve the cystic duct.

## Discussion

In this cohort of 219 patients who underwent ERCP with FCSEMS for a malignant or benign bile duct stricture, there was no association between the use of a FCSEMS with transmural side holes on rates of premature bile duct obstruction or acute cholecystitis. One potential explanation for these findings is the inherent technical limitation of side-hole design. Although side holes are intended to facilitate cystic duct drainage, their effectiveness depends on precise alignment with the cystic duct orifice, which is often difficult to determine or achieve at the time of ERCP. In this cohort, only 45% of cases with an intact gallbladder had visualization of the cystic duct during ERCP. The reasons include intentional underfilling of the bile duct and involvement of the cystic duct within the stricture. It may be counterproductive to intentionally fill the cystic duct so that side holes can be positioned optimally, since overfilling the biliary tree is associated with a higher rate of biliary adverse events.^16, 18–19^ As a result, side holes may offer an inconsistent or unreliable mechanism for preserving cystic duct drainage in routine clinical practice.

Similarly, side holes were not associated with higher rates of premature bile duct obstruction, suggesting these holes do not predispose to higher rates of tissue ingrowth – a common etiology of premature occlusion with uncovered SEMS, impact rates of spontaneous stent migration, or interfere with planned removal (there were no technical failures at stent removal).

Acute cholecystitis is a well-defined adverse event following ERCP. Since the advent of FCSEMS, there has been an ongoing concern that a coated metallic stent leads to a higher risk of acute cholecystitis by “jailing” the cystic duct insertion. However, there are other plausible etiologies for post-ERCP cholecystitis, especially in the setting of distal malignant bile duct obstruction, including tumor progression with secondary involvement of the cystic duct, immunocompromised state, and use of opioids, among others.^7^ Comparative analyses of uncovered SEMS with FCSEMS have failed to demonstrate a consistent and clinically significant difference in acute cholecystitis rates, probably because the etiology of acute cholecystitis is more often patient-related (i.e., disease occlusion of the cystic duct) or ERCP-related (bacterial translocation) and less attributable to stent-induced occlusion of the cystic duct. Our results support these hypotheses, although the number of cholecystitis events in our study were too small to allow for comparative testing.

Collectively, observations suggest that FCSEMS reduce the risk of tumor ingrowth as a cause for recurrent bile duct obstruction, but this is at least partially offset by higher rates of stent migration and clogging with debris.^1,3,6–9,20,21^ In this cohort, all patients received a FCSEMS with antimigration anchoring fins which were incorporated into the stent design to minimize rates of spontaneous migration (we observed only 2/219 (0.9%) stent migrations). We intentionally excluded patients who received an uncovered SEMS to focus on the relationship between transmural side holes and stent-associated adverse events.

This study adds to a body of literature supporting the use of FCSEMS for malignant and benign etiologies of biliary obstruction. Adverse events including cholangitis or cholecystitis remained low, supporting the ongoing use of ERCP with FCSEMS as an effective strategy for the palliation of malignant and treatment of benign biliary obstruction. The observed adverse event rates (cholecystitis: 5.4%, acute cholangitis 3.1%) are comparable to prior cohort studies: Saxena et al. found rates of cholangitis to be 4.1%, and Kogure et al. found rates of cholecystitis and acute cholangitis to be 4% each.^2,5^ From a practical standpoint, these findings suggest that side holes should not be a primary factor in FCSEMS selection. Instead, stent diameter and length remain the most important considerations. This may help simplify clinical decision-making and stent inventory without negatively affecting outcomes.

Because the overall rates of cholecystitis and biliary obstruction were low, this study may not be powered to detect small differences between groups. However, the low event rates in both groups suggest that any clinical benefit of side holes is limited. The sample size was too small to determine whether cystic duct involvement within the stricture and crossing the cystic duct were associated with premature biliary obstruction and cholecystitis. However, observed differences in rates of cholecystitis (3% when a FCSEMS doesn’t cross the cystic duct vs. 14% when it does) suggest this is a more important factor than stent type.

The principal strengths of this study include the consistent use of one FCSEMS type across the cohort, allowing for the between-group comparison of transmural side holes being present. Also, we required medical record activity to confirm follow-up time without an adverse event to minimize measurement bias. Still, the study has some limitations. Its retrospective design and non-protocolized stent selection reflect real-world clinical practice but may introduce selection bias. Importantly, a higher proportion of patients with an intact gallbladder who received a FCSEMS with side holes because of the endoscopist’s perceived concern with acute cholecystitis and benefit of this stent type. However, even when adjusting for the presence of an intact gallbladder and opacification of the cystic duct in multivariate models, the rates of biliary obstruction, and rates of acute cholecystitis, respectively, were similar between groups. Due to the retrospective study design and inconsistent visualization of the cystic duct during the index cholangiography, we cannot comment on the precise rate of cystic duct involvement at the time of FCSEMS placement, however our data indicates that stents traversed the cystic duct approximately 30-40% of the time, and at similar rates between groups. We believe it is unlikely that there were inherent differences between the two groups since the distribution of stent lengths 8cm and greater (which are the stents most likely to cross the cystic duct) were similar. There was also a higher proportion of malignant strictures in the side hole group; these patients undergo planned removal infrequently, and the malignancy may confer a higher rate of cholecystitis compared to benign etiologies. Nevertheless, after adjusting for malignant etiology of stricture in our regression model, the use of a FCSEMS with side holes was not associated with a lower rate of premature stent occlusion or acute cholecystitis as a time-to-event outcome measure which accounts for variable follow-up times.

These initial data suggest that transmural side holes may not reduce rates of premature biliary obstruction or acute cholecystitis as previously assumed, and support the design of a prospective, multi-center randomized trial stratified by malignancy and cystic duct involvement to better assess the incremental benefit of side holes. Until such data is available, our findings do not suggest stents with side holes alone reliably prevent cholangitis or cholecystitis.

## Data Availability

All data produced in the present study are available upon reasonable request to the authors

## References

1. Ghazi R, AbiMansour JP, Mahmoud T et al. Uncovered versus fully covered self-expandable metal stents for the management of distal malignant biliary obstruction. Gastrointestinal endoscopy 2023; 98: 577–584. e574

2. Kogure H, Ryozawa S, Maetani I et al. A prospective multicenter study of a fully covered metal stent in patients with distal malignant biliary obstruction: WATCH-2 study. Digestive diseases and sciences 2018; 63: 2466–2473

3. Tringali A, Hassan C, Rota M et al. Covered vs. uncovered self-expandable metal stents for malignant distal biliary strictures: a systematic review and meta-analysis. Endoscopy 2018; 50: 631–641

4. Tamura T, Yamai T, Uza N et al. Adverse events of self-expandable metal stent placement for malignant distal biliary obstruction: a large multicenter study. Gastrointestinal Endoscopy 2024; 99: 61–72. e68

5. Saxena P, Diehl DL, Kumbhari V et al. A US multicenter study of safety and efficacy of fully covered self-expandable metallic stents in benign extrahepatic biliary strictures. Digestive diseases and sciences 2015; 60: 3442–3448

6. Li J, Li T, Sun P et al. Covered versus uncovered self-expandable metal stents for managing malignant distal biliary obstruction: a meta-analysis. PloS one 2016; 11: e0149066

7. Vanella G, Coluccio C, Cucchetti A et al. Fully covered versus partially covered self-expandable metal stents for palliation of distal malignant biliary obstruction: a systematic review and meta-analysis. Gastrointestinal Endoscopy 2024; 99: 314–322. e319

8. Lam R, Muniraj T. Fully covered metal biliary stents: A review of the literature. World journal of gastroenterology 2021; 27: 6357

9. Bauss J, AbiMansour JP, Simadibrata DM et al. Comparison of fully covered self-expandable metal stents with and without anti-migration fins for the management of distal malignant biliary obstruction. Gastrointestinal endoscopy 2025.

10. Lopimpisuth C, Vedantam S, Danpanichkul P et al. Postprocedural cholecystitis following covered self-expandable metal stent placement in patients with distal malignant biliary obstruction: a systematic review and meta-analysis. Gastrointestinal endoscopy 2025.

11. Jung JH, Park SW, Hyun B et al. Identification of risk factors for obstructive cholecystitis following placement of biliary stent in unresectable malignant biliary obstruction: a 5-year retrospective analysis in single center. Surgical Endoscopy 2021; 35: 2679–2689

12. Matsumi A, Kato H, Ogawa T et al. Risk factors and treatment strategies for cholecystitis after metallic stent placement for malignant biliary obstruction: a multicenter retrospective study. Gastrointestinal endoscopy 2024; 100: 76–84

13. Ishii T, Hayashi T, Yamazaki H et al. Risk factors for early and late cholecystitis after covered metal stent placement for distal biliary obstruction. Journal of Hepato Biliary Pancreatic Sciences 2023; 30: 1180–1187

14. Chandrasekhara V, Khashab MA, Muthusamy VR et al. Adverse events associated with ERCP. Gastrointestinal endoscopy 2017; 85: 32–47

15. Kulpatcharapong S, Piyachaturawat P, Mekaroonkamol P et al. Efficacy of multi-hole self-expandable metal stent compared to fully covered and uncovered self-expandable metal stents in patients with unresectable malignant distal biliary obstruction: a propensity analysis. Surgical Endoscopy 2024; 38(1), 212–221.

16. Takahashi S, Takeda T, Kobayashi M et al. Efficacy and safety of a novel multi hole fully covered self expandable metallic stent for malignant distal biliary obstruction: Multicenter retrospective study. Digestive Endoscopy 2025; 37(7), 766–774.

17. Kiriyama S, Kozaka K, Takada T, et al. Tokyo Guidelines 2018: diagnostic criteria and severity grading of acute cholangitis (with videos). Journal of Hepato Biliary Pancreatic Sciences 2018; 25: 17–30

18. Talukdar R. Complications of ERCP. Best practice & research Clinical gastroenterology 2016; 30: 793–805

19. Cotton PB, Connor P, Rawls E et al. Infection after ERCP, and antibiotic prophylaxis: a sequential quality-improvement approach over 11 years. Gastrointestinal endoscopy 2008; 67: 471–475

20. Conio M, Mangiavillano B, Caruso A et al. Covered versus uncovered self-expandable metal stent for palliation of primary malignant extrahepatic biliary strictures: a randomized multicenter study. Gastrointestinal Endoscopy 2018; 88: 283–291. e283

21. Conigliaro R, Pigò F, Bertani H et al. Migration rate using fully covered metal stent in anastomotic strictures after liver transplantation: Results from the BASALT study group. Liver International 2022; 42: 1861–1871

